# An empirical estimate of the infection fatality rate of COVID-19 from the first Italian outbreak

**DOI:** 10.1101/2020.04.18.20070912

**Authors:** Gianluca Rinaldi, Matteo Paradisi

## Abstract

**Background:** The coronavirus 2019 (COVID-19) pandemic has been spreading globally for months, yet the infection fatality ratio of the disease is still uncertain. This is partly because of inconsistencies in testing and death reporting standards across countries. We provide estimates which don’t rely on official cases and deaths data but only on population level statistics.

**Methods:** We collected demographic and death records data from the Italian Institute of Statistics. We focus on the area in Italy that experienced the initial outbreak of COVID-19 and estimated a Bayesian model fitting age-stratified mortality data from 2020 and previous years. We also assessed the sensitivity of our results to alternative assumptions on the proportion of population infected. Based on our estimates we finally studied the heterogeneity in overall lethality across countries.

**Findings:** We estimate an overall infection fatality rate of 1.31% (95% credible interval [CrI] 0.94 – 1.89), as well as large differences by age, with a low infection fatality rate of 0.05% for under 60 year old (CrI 0 – 0.17) and a substantially higher 4.16% (CrI 3.05 – 5.80) for people above 60 years of age. In our sensitivity analysis, we found that even under extreme assumptions, our method delivered useful information. For instance, even if only 10% of the population were infected, the infection fatality rate would not rise above 0.2% for people under 60. Finally, using data on demographics we show large expected heterogeneity in overall IFR across countries.

**Interpretation:** Our empirical estimates show a sharp difference in fatality rates between young and old people and rule out overall fatality ratios below 0.5% in populations with more than 30% over 60 years old.

## 1. Introduction

Estimating the severity of the coronavirus disease 2019 (COVID-19), caused by the novel severe acute respiratory syndrome coronavirus 2 (SARS-CoV-2) across demographic groups is of medical interest, but will also be crucial in the design of health and economic policies to deal with the next phases of the pandemic.

Several approaches to measure lethality have been proposed since the start of the outbreak ^1,2,3,4^, with most of the relevant population infected. Given the mounting evidence that a substantial proportion of infected people are either asymptomatic or display very mild symptoms^5,6^, as well as the difficulties many countries are encountering in ramping up testing, it has been difficult to obtain precise estimates of the total number of infected. Testing mainly symptomatic cases on the basis of clinical studies on the symptoms of COVID-19^7,8,9,10,11^, as announced for instance by the Italian government, might also have led to underestimating the total number of infected. While measurement of deaths is reliable, statistically significant deviations in total deaths relative to previous years have been observed in the most affected areas, leading to concerns that official COVID-19 death counts might also be underestimated in some cases^12^. Together, those hurdles make estimating the true infection fatality rate challenging.

To sidestep some of those issues, in this article we used an empirical approach employing publicly available aggregate deaths and demographic data to obtain infection fatality ratio estimates without relying on official data on COVID-19 positive cases and deaths. The key observation is that, assuming an accurate measurement of fatalities in a population, infection fatality ratio estimates are less strongly dependent on accurate measurement of total cases when the share of population infected is larger: keeping the number of deaths fixed, the estimate changes much less when the fraction of the population infected varies from 40% to 60% than when it changes from 2% to 3%, simply because ratios are nonlinear.

We were therefore able to obtain precise fatality estimates by age range focusing on one of the hardest hit areas in Lombardy, which was placed under lock-down order already on February 21st 2020. This area includes ten towns and has a population of around 50 thousands people. The first recorded patient infected through community spread in Italy was admitted to intensive care on February 20th and the area already had 36 confirmed cases the day after^13^. While widespread randomized antibody testing hasn’t yet been conducted in this area, 30% of a sample of blood donors from all ten municipalities tested positive to antibody^14^, and a smaller sample of 60 asymptomatic blood donors in one of the towns under lock-down showed 40 (66%) positive cases^15^. Although these samples might not be fully representative of the population of these areas, this figure is highly suggestive of a widespread contagion.

## 2. Methodology

### 2.1. Data

We focused on ten Italian municipalities in Lombardy that experienced the initial outbreak of COVID-19. Data on deaths has been collected from the Italian Institute of Statistics (ISTAT)^16^. We built estimates of total death counts based on daily data on recorded deaths for 2020 and previous years for the period 2015-2019 that ISTAT collects from the *Anagrafe Nazionale della Popolazione Residente* (National Census of Resident Population). The data contains information about gender and age group (in 5-years bins) for each recorded death until April 15th 2020. ISTAT has released data for only eight of the ten municipalities that experienced the initial outbreak until April 15th, while information on nine municipalities is available until April 4th. Hence, we focused our baseline analysis on the latter subsample, while using the smaller sample until April 15th to test the robustness of our results. The excluded town in the baseline sample is much smaller than the others: its total population in 2019 was 1,118, while the remaining nine municipalities had a total population of 49,431. For one of the municipalities (Casalpusterlengo) we used the most recent data from^16^ available until May 31st, while we drew information for the period April 1st to April 4th from a previous data release^17^.

Death counts data has been complemented with information provided by ISTAT on the demographics of each municipality^18^. For every city we collected total population by age range in each year from 2015 to 2019. We used year 2019 information for year 2020 since data on 2020 has not been released yet. The municipality of Castelgerundo was created in 2018 by the union of the municipalities of Camairago and Cavacurta. We therefore used total population counts from the latter two for the years 2015-2018.

Finally, data on demographics for OECD countries was collected^19^. Unfortunately, OECD age groups do not perfectly overlap with the age bins in the Italian death data. Indeed, there is a one year difference: OECD age groups are, for instance, 20 – 24, while the Italian deaths data uses bins such as 21 – 25. Since we aggregate across multiple bins this is not a major concern, for instance the 21 – 40 deaths data bin corresponds to 20 – 39 in the OECD data.

### 2.2. Comparing 2020 deaths to previous years

In order to motivate the use of administrative death counts for our infection fatality rate estimation, we begin by analysing patterns in overall mortality in 2020 and previous years. In particular, on every day between February 21st (the beginning of the outbreak) and April 4th the difference between the number of 2020 deaths and the 2015-2019 average has been computed for total population and for different age groups. We compared total deaths to average number and fluctuations in deaths in past years to assess the signal to noise ratio of this measure.

### 2.3. Bayesian Estimation of COVID-19 infection fatality rate

We employ a Bayesian framework to estimate the infection fatality rate of COVID-19 by adapting a standard binomial mortality model to our setting^20^. The likelihood function of the model is obtained by assuming deaths in the period between February 28th and April 4th of each year are binomially distributed according to:

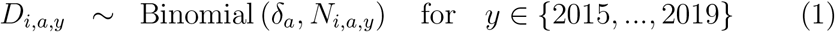

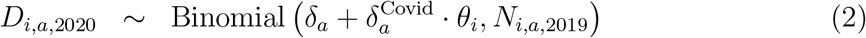

where *i* denotes the municipality, y the year, and *a* the age range. We used seven age ranges: 0 – 20, 21 – 40, 41 – 50, 51 – 60, 61 – 70, 71 – 80, 81+. *D_i,a,y_* and *N_i,a,y_* are the total deaths and population in town *i*, year *y* and age range *a,* respectively. For population in 2020, we use 2019 data since 2020 data is not yet available. The baseline lethality rates *δ_a_* are heterogeneous across age ranges, but were assumed to be constant across municipalities and years.

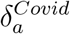 is the infection fatality rate for age range *a* and we assumed 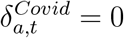 in every year before 2020 when COVID-19 was not present. We also assumed that infection rates *θ_i_* are heterogeneous across municipalities but constant across age groups.

We assumed the following priors for the parameters of interest:

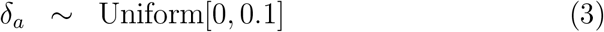

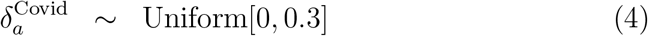

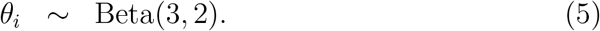

Priors on baseline and COVID-19 death rates were chosen to be uninformative, while we chose the prior on infection rates to reflect the results of the antibody testing in one of the municipalities^15^, while at the same time maintaining a weakly informative prior.

We implemented a Bayesian procedure to derive point estimates and credible intervals for the infection fatality rates. The model was estimated using Markov Chain Monte Carlo (MCMC). We calculated the median and 95% credible interval using to the quantiles of the posterior distribution for all parameters. To check the sensitivity of our estimates we also calculated point estimates as the mode of the posterior distribution and confidence intervals as 95% highest posterior density interval^21^.

We fitted our model using R version 3.6.2. We drew 100,000 samples from the joint posterior distribution and used 10 independent chains, discarding the first 1000 samples for each chain. Trace plots of the Markov Chain Monte Carlo as well as posterior distributions for each variable are reported in Appendix A. All analyses are fully reproducible with the code available online.

Because our Bayesian procedures relies on modelling assumptions to derive the infected portion of the population, we also implemented a more agnostic approach, showing how infection fatality rates vary by contagion rate. In this exercise we computed the infection fatality rates from a simplified version of the model above in which we set a degenerate prior for each of the 6_i_ to be equal to a constant in the interval [0.1,1] and estimated the model for each choice.

### 2.4. Projections of IFR Across Countries

We used the estimates of our Bayesian model to draw projections on the overall lethality in OECD countries. We built these projections assuming homogeneous contagion across age groups and using each country’s demographic composition to re-weight our IFR estimates through linear combinations. Point estimates in this exercise were produced combining the medians of the posteriors of different age groups, while 95% credible intervals were obtained with linear combinations of the extremes of the 95% credible intervals of the age-specific IFRs.

## 3. Results

### 3.1. Raw Deaths Counts

We documented a substantial increase in total deaths at the beginning of the outbreak. Figure 1 shows the total daily deaths counts in the nine municipalities for 2020 and 2015-2019 (average) in the period between Jan 1st and April 4th. The year 2020 and previous years are very similar preceding the last week of February, when the first COVID-19 cases have been discovered. Starting that week, we observed a spike in the number of deaths. This spike is clearly related to COVID-19 as it starts at the beginning of the outbreak and it significantly overcomes average fluctuations in deaths observed in previous years. In total, deaths over the period from February 21st to April 4th in 2020 were almost five times the average in previous years over the same period of time (371 vs 73).

**Figure 1:**
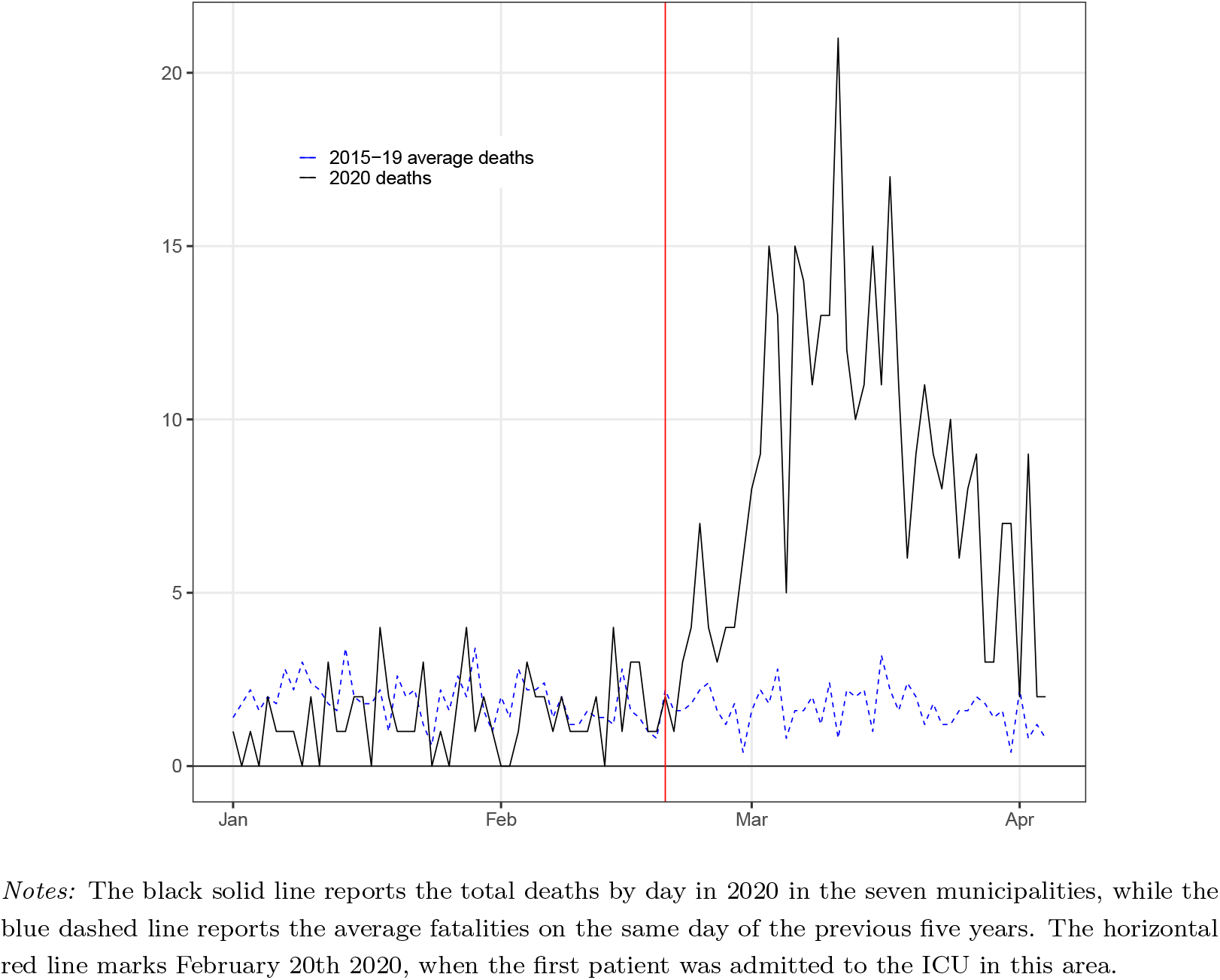
Daily deaths in 2020 and previous years average

### 3.2. Infection Fatality Ratio Estimates

Using our Bayesian model we estimated an overall infection fatality ratio of 1.31% (95% credible interval [CrI] 0.94–1.89). We also uncovered substantial heterogeneity by age. For under 60 years old the infection fatality rate was 0.05% (CrI 0 – 0.17, Table 2), while for over 60 years old it was 4.16% (CrI 3.05– 5.80, Table 2).

**Table 1:** Total population and deaths by age range

| Age Range | Total Population |  |  |  |  | Total Deaths |  |  |  |  |  |
| --- | --- | --- | --- | --- | --- | --- | --- | --- | --- | --- | --- |
|  | 2015 | 2016 | 2017 | 2018 | 2019 | 2015 | 2016 | 2017 | 2018 | 2019 | 2020 |
| 0 – 20 | 8452 | 8447 | 8420 | 8438 | 8460 | 0 | 0 | 0 | 0 | 0 | 1 |
| 21 – 40 | 10777 | 10616 | 10374 | 10233 | 10217 | 0 | 0 | 1 | 0 | 0 | 0 |
| 41 – 50 | 8407 | 8239 | 8053 | 7884 | 7630 | 1 | 2 | 0 | 1 | 2 | 2 |
| 51 – 60 | 7258 | 7480 | 7646 | 7872 | 7992 | 3 | 2 | 5 | 2 | 5 | 6 |
| 61 – 70 | 6037 | 6024 | 6022 | 5984 | 5984 | 5 | 5 | 4 | 9 | 7 | 31 |
| 71 – 80 | 4943 | 4968 | 5054 | 5090 | 5144 | 13 | 10 | 17 | 18 | 10 | 120 |
| 81+ | 3556 | 3607 | 3732 | 3822 | 4004 | 57 | 36 | 43 | 48 | 59 | 211 |
| Overall | 49430 | 49381 | 49301 | 49323 | 49431 | 79 | 55 | 70 | 78 | 83 | 371 |

*Notes:* This table reports descriptive statistics by age range on total population for 2015-2019, and deaths for 2015-2020. Data refers to the period from January 1st to April 4th.

**Table 2:**
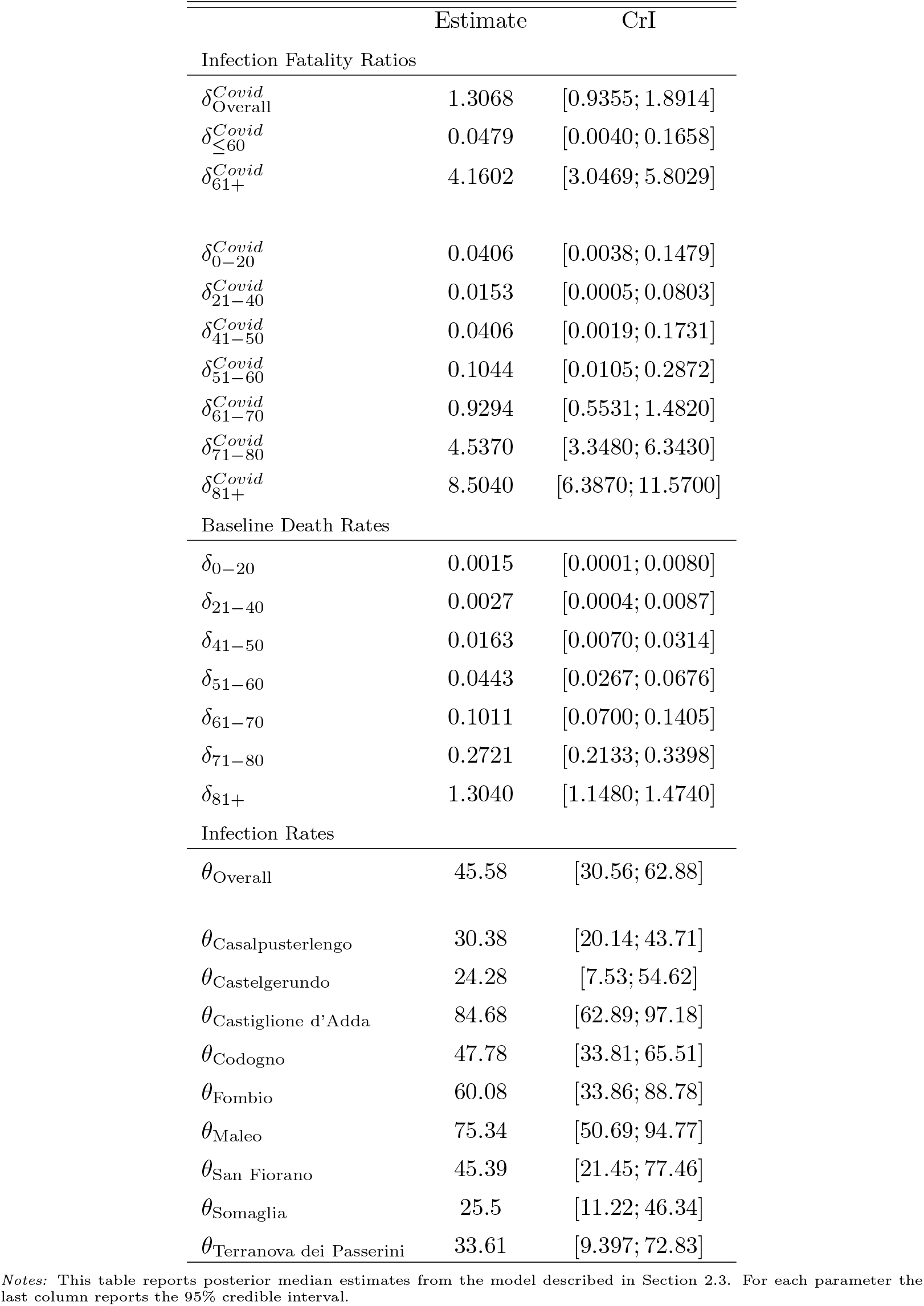
Model Estimates

Figure 2 shows the estimated infection fatality ratios by age group together with 95% credible intervals and interquartile range. As expected, infection fatality ratios were found much larger for older age groups. Point estimates are 4.54 and 8.50 for 71-80 and 81+, respectively. We cannot exclude, however, that the infection fatality ratio for over 80 years old is as high as 11. 57% or as low as 6.39%. Interestingly, we found an infection fatality rate close to zero for under 50 years old, and around 0.1% for the 51-60 group. For robustness, we have also recalculated all point estimates using the mode and confidence intervals as the highest posterior density interval. Results remained virtually identical (data not shown). Moreover, we checked that results remained unchanged when extending the period of analysis until April 15th and looking at the eight municipalities for which we have data until this date (data not shown).

**Figure 2:**
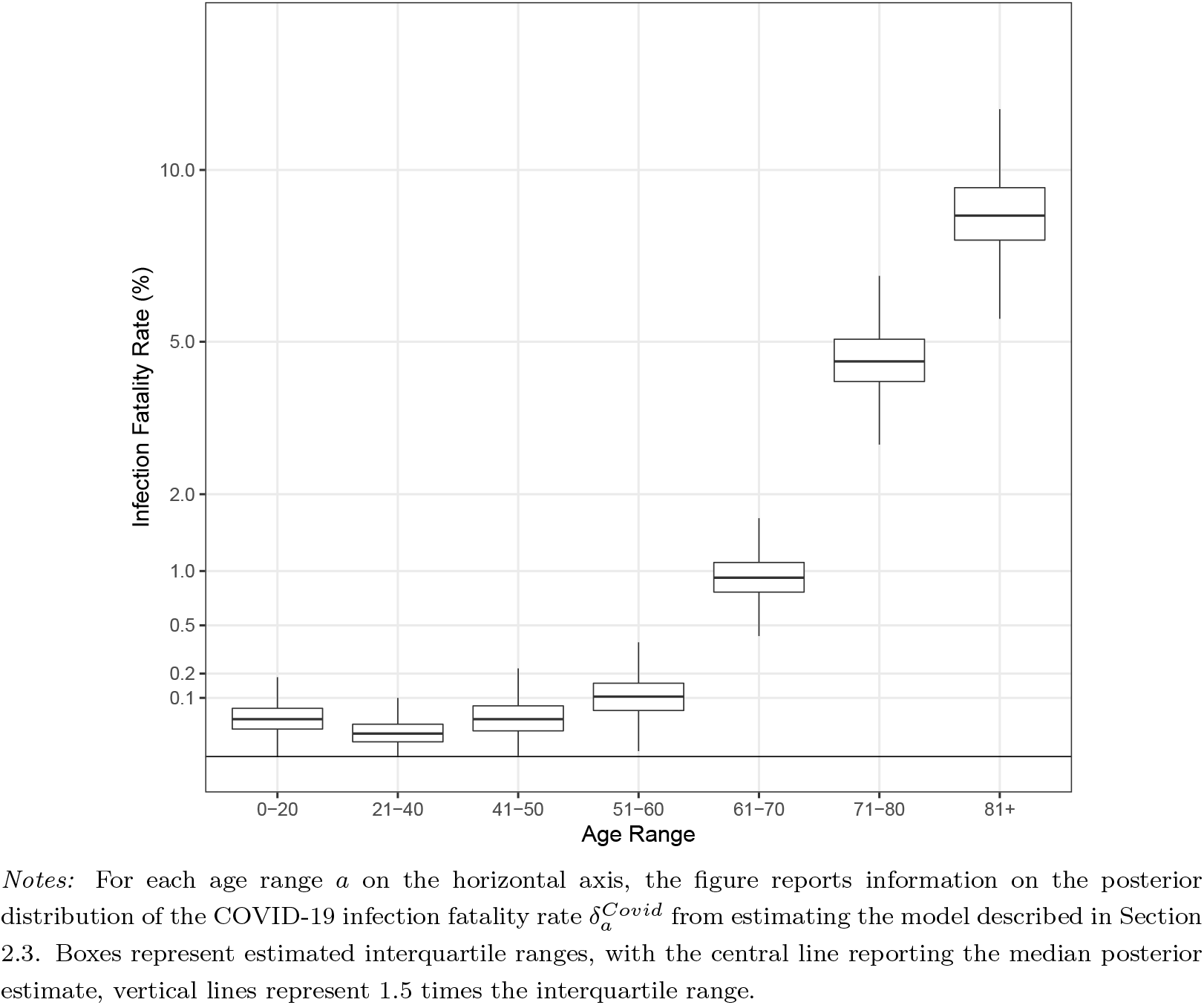
Estimates of the infection fatality rate by age range

Estimated infections rates were also heterogeneous by town, ranging between 24.3% and 84.7% (Table 2). Interestingly, Castiglione d’Adda, where antibody tests conducted on a sample of individuals detected a 66.6% infection rate, resulted as the municipality with the largest share of the population infected (84.7%). We estimate a population weighted overall infection rate for the nine towns of 45.58% (CrI 30.56% – 62.88%). This is broadly consistent with a recent study on blood donors for the entire area^14^ has found a 30% overall infection rate.

We then performed an exercise to assess how sensitive our infection fatality rate estimates are to different levels of contagion (Figure 3). Focusing on a large range of potential infection rates, we found that even in the conservative assumption that only 15% of the population was infected, under 60 years old still experienced an infection fatality rate significantly below 1%, with under 40 being around 0.1%. Obviously, estimates spiked for older age groups as we set the infection rate very low. These results confirm the view that the infection has low lethality rates for younger individuals, but large rates for the elders. Moreover, this exercise showed that the overall infection fatality rate was significantly above zero and around 0. 5% even in the most conservative assumption of 100% contagion.

**Figure 3:**
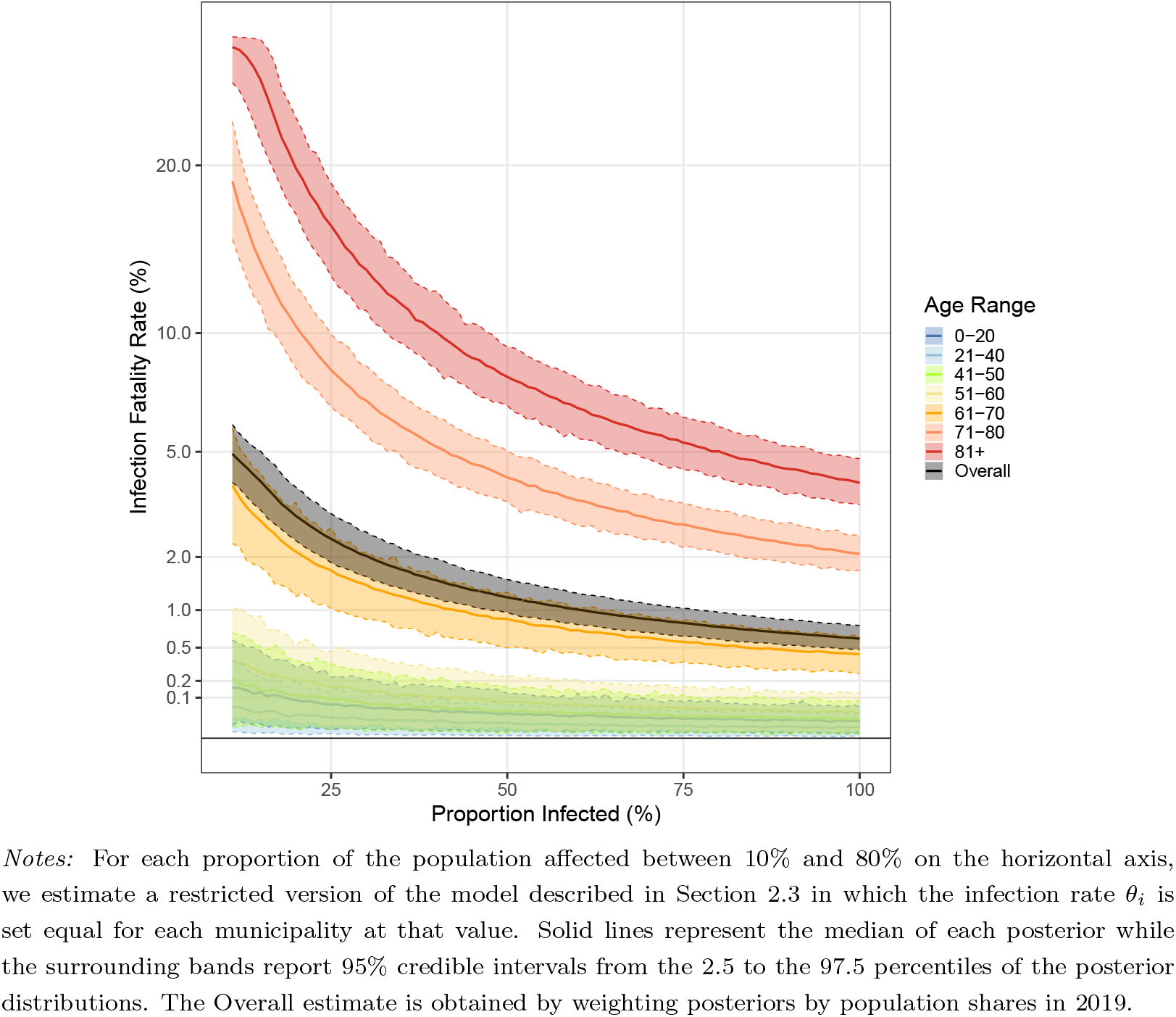
Estimates of infection fatality rate by age and proportion of population infected

Finally, we used our estimates to build overall IFRs for OECD countries. We re-weighted the IFR of each age group by its respective share of the total population of each country. This procedure relies on the assumption that contagion was homogeneous across age groups. Figure 4 shows the results ranking countries based on their projected IFR. Given their high shares of over 60 years old, Japan and Italy are the countries with the highest IFR, with point estimates of 1.44% and 1.19%, respectively. The Europe-28 IFR is slightly above 1%. Interestingly, the U.S. are projected to have a IFR below 1% and equal to 0.8% if contagion spread homogeneously across age groups. Very few developed countries are expected to have a IFR below 0.5% in the scenario of homogeneous infection across ages.

**Figure 4:**
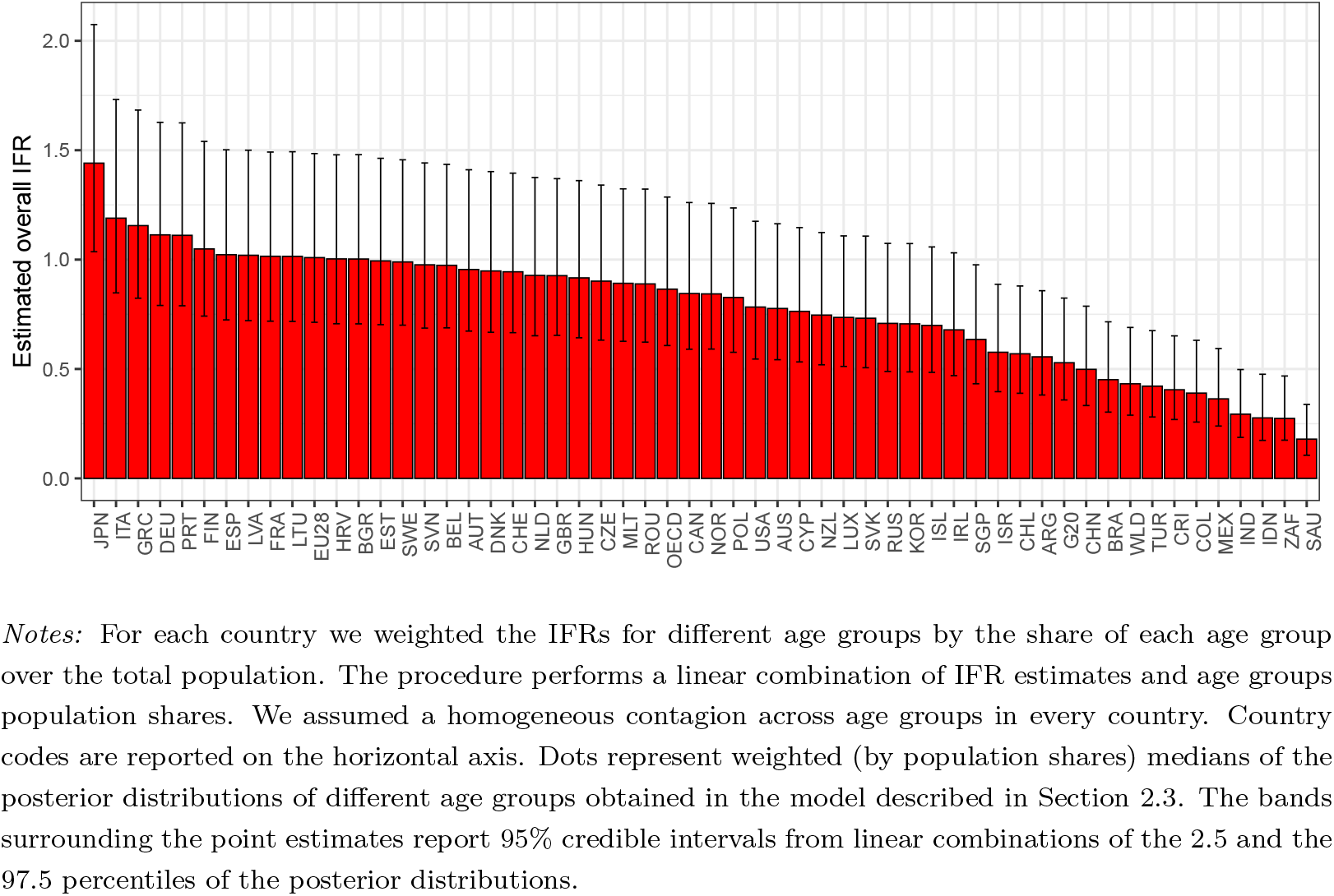
Estimates of infection fatality rate by country

## 4. Discussion

In this paper we estimated the infection fatality rate of COVID-19 from administrative death counts on nine Italian municipalities that experienced the first outbreak of this desease in late February 2020. We found an overall infection fatality rate of 1.31% (CrI 0.94 – 1.89). We uncovered significant heterogeneity across age groups. Under 60 years old have infection fatality ratios around 0.05% (CrI 0.00 – 0.17). On the contrary, older people are at significantly larger risk: over-60 infection fatality ratio is 4.16% (CrI 3.05 – 5.80), and for over-80 it is estimated at 8.50% (CrI 6.4 – 11.57). We excluded very low fatality rates even under very conservative assumptions on the population infection rate: if the entire population had been infected, the overall infection fatality ratio would still be around 0.5%. Finally, we showed projections of overall IFRs across countries in the scenario of equal contagion across age groups. Japan and Italy are the countries expected to suffer the most with IFRs of 1.44% and 1.19%, while the US have a smaller IFR equal to about 0.8%.

Our result for overall infection fatality ratio is larger than estimates in^22^ based on travellers’ data. It is, however, remarkably close to estimates for two case studies where the entire population was tested: Diamond Princess Cruise (1.3%)^23^, and Vo’ Euganeo (1%) - a 3,000 inhabitants municipality in the Italian Veneto region. Despite the similarity there are two main reasons to expect differences between our estimates and those in the two mentioned case studies. First, we estimate a large infection fatality ratio for over 80 years old who are likely underrepresented in the Diamond Princess Cruise. Second, total population in both Diamond Princess Cruise and Vo’ Euganeo is limited and for this reason those estimates might be affected by undersampling of the number of infected in the tails. These small differences notwithstanding, all these figures are substantially lower than estimated case fatality rates (CFRs) computed with official contagion data and for previous influenza pandemics^24^. The most plausible reason is that COVID-19 case counts are subject to downward biases due to limited testing capacity and testing strategies that prioritize symptomatic cases despite a large number of asymptomatic patients.

Our estimates could suffer from the fact that deaths data is missing for the period after April 4th 2020. This would be an issue if the contagion had not stopped by that date and therefore useful information on deaths could not be used in our model. However, as the trend in number of deaths suggests, the contagion likely stopped by April 4th in these nine municipalities. Indeed, the number of deaths in the last days of our sample went back to the average number in the previous five years. Moreover, using data available until April 15th for eight of the nine municipalities, we checked that the number of deaths remained aligned to the average in previous years between April 4th and April 15th. For this reason, there are reasons to believe that data for the following weeks would not be very informative on the lethality of the first wave of contagion.

One limitation of our estimates is that they should not be taken at face value in the analysis of contexts where the hospital system is under stress or close to capacity. This is because our exercise was performed on a case study at the beginning of the Italian outbreak when the hospital system was still fully functioning. Moreover, the quarantine measures implemented in the nine municipalities on February 21st likely reduced contagion, potentially affecting infection fatality ratios and making them hard to extrapolate to context where similar measures were not undertaken.

Another limitation is that our model assumed a constant baseline lethality rate in absence of COVID-19. This implies that the COVID-19 outbreak did not change the baseline death rate in the population. Plausibly, the lockdown policy decreased deaths from, among others, violence and traffic, while at the same time the outbreak could have increased other fatalities due to lower availability of healthcare resources for other diseases. Fluctuations due to these causes are, however, likely to be quantitatively small compared to the large spike in deaths that we observed in 2020, where total deaths were five times the average in previous years over the same period of time (371 vs 73). For this reason, confounding factors in baseline deaths levels should not have substantially affected the signal to noise ratio of our death data.

In conclusion, our results support the need for isolating policies especially for the elder part of the population given the extremely high fatality rates from COVID-19 that we estimated.

## Contributors

GR conceived the study with input from MP. MP collected demographic and deaths data and cross checked the accuracy of the data. GR developed the code for the Bayesian model and estimated it with input from MP. MP produced the first draft of the manuscript. All authors contributed to the final draft.

## Declaration of interests

GR and MP declare no competing interests.

## Data Sharing

All data is publicly available and can be found at the cited sources. Code for replication is available at https://github.com/gianlucaRinaldi/covid_IFR_Lombardy.

## Data Availability

All data is publicly available and can be found at the cited sources.

https://github.com/gianlucaRinaldi/covid_IFR_Lombardy

## Appendix A. Markov Chain Monte Carlo Estimation diagnostics

**Figure Appendix A.1:**
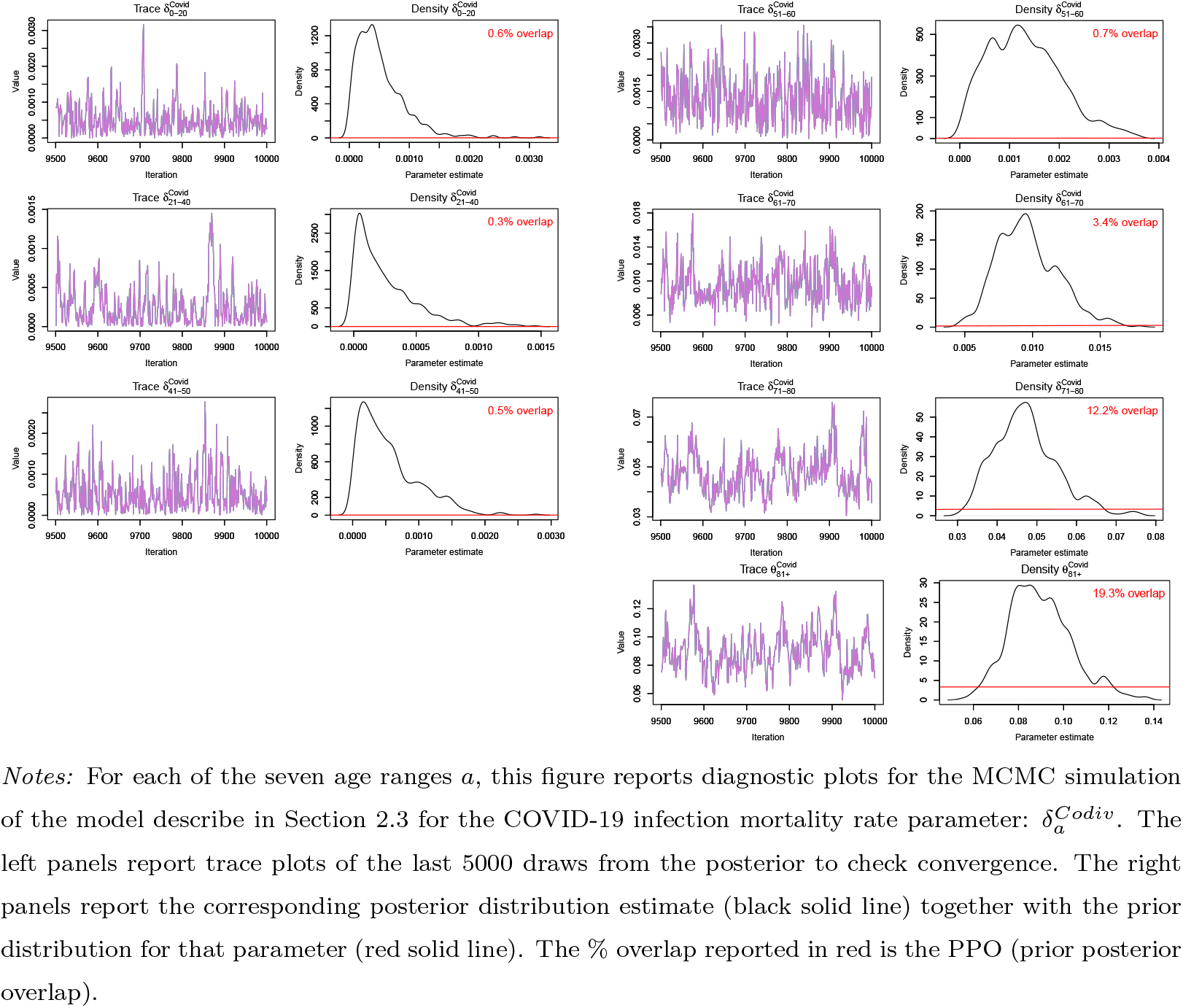
Trace and density plots for MCMC posteriors of 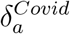 for each age range

**Figure Appendix A.2:**
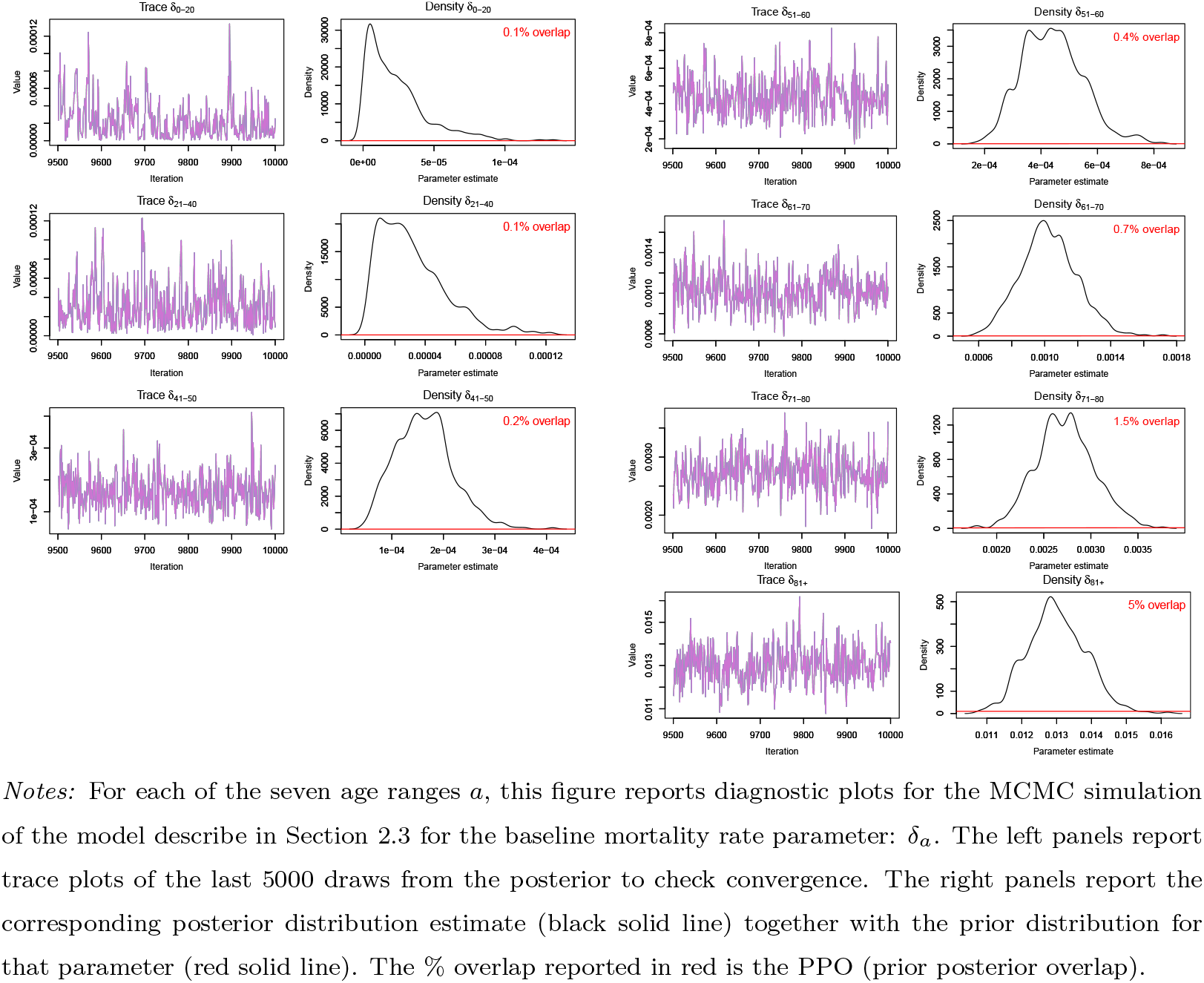
Trace and density plots for MCMC posteriors of *δ_a_* for each age range

**Figure Appendix A.3:**
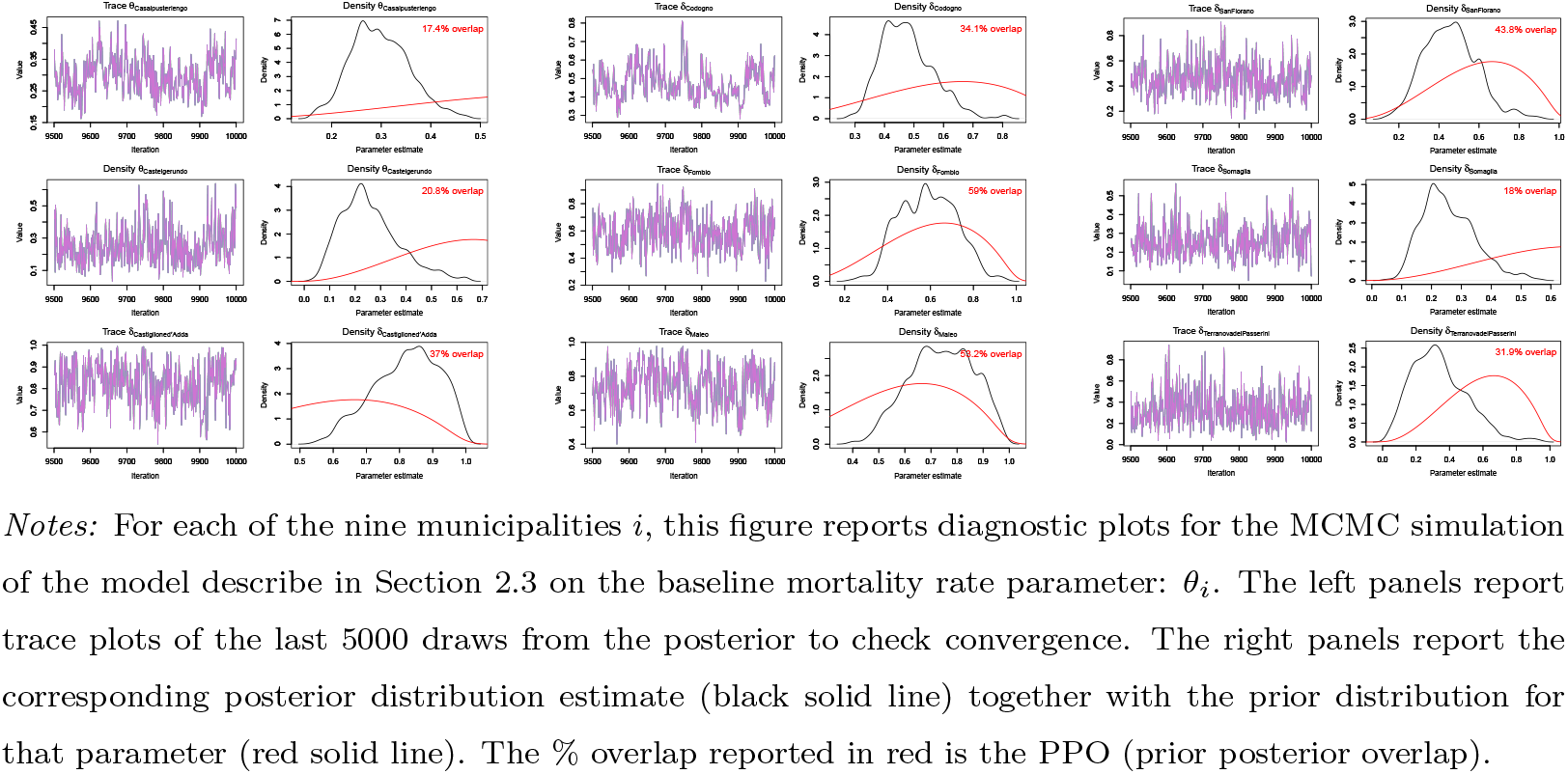
Trace and density plots for MCMC posteriors of *θ_i_* each municipality

## Notes

### Competing Interest Statement

The authors have declared no competing interest.

### Funding Statement

The funders had no role in study design, data collection, data analysis, data interpretation, or writing of the report. All authors had full access to all data in the study and had final responsibility for the decision to submit for publication.

